# Modeling COVID-19 Growing Trends to Reveal the Differences in the Effectiveness of Non-Pharmaceutical Interventions among Countries in the World

**DOI:** 10.1101/2020.04.22.20075846

**Authors:** You Chen, Yubo Feng, Chao Yan, Xinmeng Zhang, Cheng Gao

## Abstract

**Objective:** We hypothesize that COVID-19 case growth data reveals the efficacy of NPIs. In this study, we conduct a secondary analysis of COVID-19 case growth data to compare the differences in the effectiveness of NPIs among 16 representative countries in the world.

**Methods:** This study leverages publicly available data to learn patterns of dynamic changes in the reproduction rate for sixteen countries covering Asia, Europe, North America, South America, Australia, and Africa. Furthermore, we model the relationships between the cumulative number of cases and the dynamic reproduction rate to characterize the effectiveness of the NPIs. We learn four levels of NPIs according to their effects in the control of COVID-19 growth and categorize the 16 countries into the corresponding groups.

**Results:** The dynamic changes of the reproduction rate are learned via linear regression models for all of the studied countries, with the average adjusted R-squared at 0.96 and the 95% confidence interval as [0.94 0.98]. China, South Korea, Argentina, and Australia are at the first level of NPIs, which are the most effective. Japan and Egypt are at the second level of NPIs, and Italy, Germany, France, Netherlands, and Spain, are at the third level. The US and UK have the most inefficient NPIs, and they are at the fourth level of NPIs.

**Conclusions:** COVID-19 case growth data provides evidence to demonstrate the effectiveness of the NPIs. Understanding the differences in the efficacy of the NPIs among countries in the world can give guidance for emergent public health events.

## Introduction

Severe acute respiratory syndrome coronavirus 2 (SARS-CoV-2) or coronavirus disease 2019 (COVID-2019), spread rapidly and globally^1-2^. The COVID-19 pandemic that was declared on March 11, 2020, has affected countries on all continents^3^. The current outbreak of COVID-19 has raised great concern worldwide^1-3^. United States of America (US) and countries (e.g., Italy, Spain, France, Germany, Netherlands, and United Kingdom (UK)) in Europe are now the epicenter of the COVID-19 pandemic^4-7^. A briefing document produced by Public Health England for the government states that the COVID-19 outbreak is expected to last around one year (until spring 2021), with around 80% of the population infected and up to 15% of people (7.9 million) requiring hospitalization in the UK^6^.

Many countries have implemented flight restrictions to others, imposed lockdowns, and closed borders to mitigate the exponential spread of COVID-19^8-9^. Within each country, many non-pharmaceutical interventions (NPIs) to reduce COVID-19 spread rates have been implemented, including social distancing, case isolation, home quarantine, school, and university close, and et ac.^9-11^.

It has been recognized that NPIs can reduce the reproduction rate, and thus change COVID-19 growth trend^9-11^. Since there are various types of NPIs, and many other factors that may impact each of them, it is very challenging to measure the impacts of NPIs on the controlling of COVID-19 growths directly. We assume that the time-series cumulative number of cases should reflect the effectiveness of the NPIs. Therefore, we propose to develop an informatics strategy to conduct secondary time-series data analysis to learn COVID-19 growing patterns to characterize the efficacy of the NPIs. We aim to model the dynamic changes in the reproduction rate over time, which will be leveraged to demonstrate the extent to which NPIs impact COVID-19 growth. Upon the relationships between the cumulative number of cases and the dynamic changes in the reproduction rate, we compare the differences in the effectiveness of NPIs across sixteen countries covering Asia, Europe, North America, South America, Australia, and Africa.

## Methods

### Study Materials

This study leverages the cumulative number of COVID-19 cases of 16 countries starting from January 22 to April 20, 2020, to learn COVID-19 growth patterns. The data is updated daily by Humanitarian Data Exchange, contributed by Johns Hopkins School of Public Health^15^.

By April 20, 2020, we collected 1,424 records, each of which represents the number of cumulative cases a country had on a specific day. For instance, the US had 759,786 cases on April 20, 2020.

### Study Design

#### Cohort creation and normalization

In preparation for this study, we composed a cohort by defining observations. An observation is a record made up of <country, date, cumulative number of confirmed cases>, as mentioned above. Since the start dates of COVID-19 pandemic for countries are different, we use the time when the 100^th^ case was confirmed as relative start dates to align COVID-19 growths for the 16 countries. Since each country has a different population (e.g., Spain – 46,754,778, and China – 1,439,323,776), we normalize the cumulative number of cases as the number of cases per million people.

### Modeling dynamic changes in the reproduction rate

We introduce three notations: i) ***d***, the number of days from the date when the 100^th^ case was confirmed; ii) ***N***_***case_d***_, the observed cumulative number of cases per million on day ***d***; and iii) ***b***, the observed cumulative number of cases per million on day 1 (the 100^th^ case was confirmed). We use the above defined three notations to model ***R***_***d***_, which is defined as the overall reproduction rate in the past ***d*** days

We assume ***N***_***case_d***_ ***= b× R***_***1***_ ^***d-1***^,***(d>1)***, give none of NPIs is adopted. Without NPIs, the COVID-19 should follow a very natural spread, which satisfies ***b× R***_***1***_ ^***d-1***^. ***R***_***1***_ is the reproduction rate on day 1. For instance, if COVID-19 infected 100 people on day 1, and each person can affect 0.5 (***R***_***1***_ is 1.5) person on average next day, then there will be 100×0.5 person infected on day 2, and the total number of infected people will be 100× (1+0.5) on day 2, and 100× (1+0.5)^2^ on day 3, and so on.

We assume ***R***_***d***_ is not fixed to be ***R***_***1***,_ and it should dynamically change over the days with NPIs adopted appropriately by a country. We use ***N***_***case_d+1***_ ***= b× R***_***d***_ ^***d***^,***(d*≥*1)*** to calculate the overall reproduction rate in the past ***d*** days - ***R***_***d***_. With the continuous NPIs, we assume ***R***_***d***_ should decrease linearly over days. We propose to leverage linear regression models to learn relationships between variables ***d*** and ***R***_***d***_ as ***R***_***d***_ = *α – β ×* ***d***. Upon the regression model, we can predict ***R***_***d***_ and ***N***_***case_d***_ on day ***d***, which we call estimated values and refer them as ***R’***_***d***_ and ***N’***_***case_d***,_ respectively.

### Learning relationships between dynamic changes in the reproduction rate and the cumulative number of cases

**Figure 1** shows an example of the relationship between day ***d*** and the cumulative number of cases ***N***_***case_d***_ on day d, which is calculated based on the ***R***_***d***_. Each point (e.g., (19,33) or (39, 33)) represents the cumulative number of cases per million population on a specific day ***d*** given the overall reproduction rate in the past ***d*** day (***R***_***d***_). The value of ***R***_***d***_ decreases over days. The point (19, 33) has higher ***R***_***d***_ than the point (39, 33), which demonstrates that the more effective NPIs (lower reproduction rate), the many more days required to reach the same cumulative number of cases. For instance, it requires 19 days to reach 33 cases per million population at a reproduction rate ***R***_***d***_ as ***1***.***3***, while 39 days to reach the same number of cases at a reproduction rate as ***1***.***2***. Leveraging more effective NPIs will require many more days to reach a targeted cumulative number of cases but, at the same time, can avoid arriving at a peak (e.g., (29, 65), as shown in **Figure 1**. From **Figure 1**, we can see that the orange line requires 20 (39 vs. 19) more days to reach the target number 33 than the blue line, while it can avoid reaching a higher peak (65 cases per million on day 29 as shown in Figure 1).

**Fig. 1.**
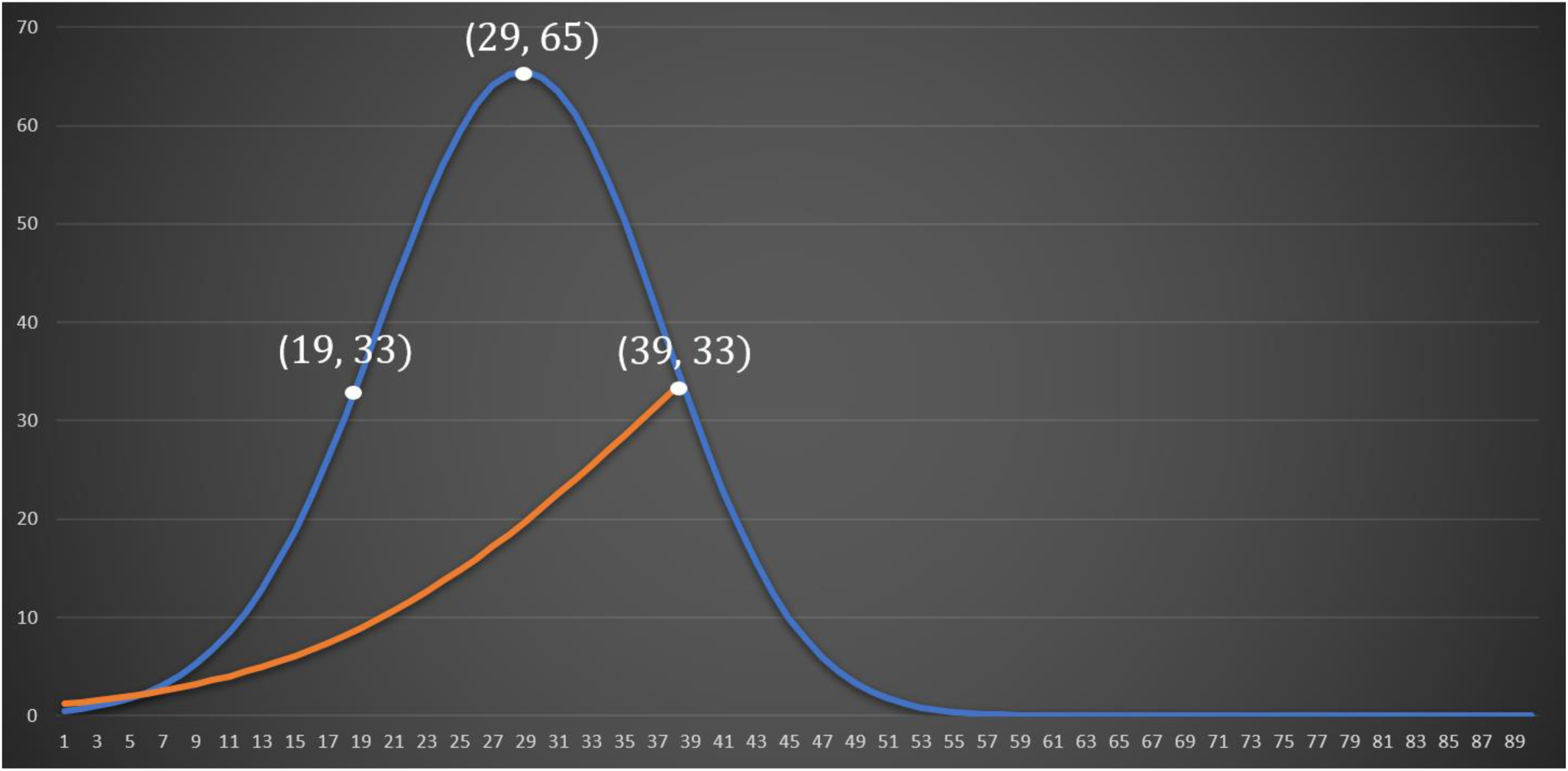
An example graph of the relationship between day ***d*** (X-axis) and the cumulative number of cases ***N***_***case_d***_ (Y-axis) on day ***d***. The figure is plotted based on the overall reproduction rate in the past ***d*** days ***R***_***d***_. Each point in the graph represents the cumulative number of cases per million population on a specific day. For instance, (19,33) shows there are 33 cases per million on day 19. **R**_**d**_ decreases over the days. The points (19,33) and (39,33) have the same target number of cases; while the number of days to arrive at the target number is different (19 vs. 39). Using more effective non-pharmaceutical interventions will make **R**_**d**_ smaller and require many more days to reach the target number. The benefit is the growth trend can avoid arriving at a higher peak ((29, 65), as shown in the figure).

We hypothesize that the effectiveness of the NPIs can be characterized by the height of the peak, which is defined as a point made up of day ***d*** and the cumulative number of cases ***N***_***case_d***_ on day ***d***. The smaller values of ***N***_***case_d***_ and ***d*** at the peak, the more effective of NPIs. According to the cumulative number of cases at the peak and the number of days reaching the peak, we developed four levels of NPIs. The four levels of NPIs are: i) the lower cumulative number of cases and the smaller number of days; ii) the lower cumulative number of cases and the bigger number of days; iii) the higher cumulative number of cases and the smaller number of days and iv) the higher cumulative number of days and the bigger number of days. Countries at the first level have the most effective NPIs (the lower cumulative number of cases and a shorter time to arrive at the peak). Countries at the fourth level have the most ineffective NPIs (the higher cumulative number of cases and a longer time to arrive at the peak).

Since some countries have not yet arrived at their peaks, we use estimated ***R’***_***d***_ to calculate the cumulative number of cases ***N’***_***case_d***,_ at the peak, and the number of days needed to reach the peak. According to ***N’***_***case_d+1***_ ***= b× R’***_***d***_ ^***d***^,***(d*≥*1)***, when the ***R’***_***d***_ decreases to a specific value ***R’***_***peak***_, the value of ***N’***_***case_d+1***_ will decrease. We set the day ***d***_***peak***_ when ***N’***_***case_d+1***_ reaches the maximum number as the turning point ((29, 65), as shown in **Figure 1**). Using the ***d***_***peak***_, we can estimate the cumulative number of cases of a country on that day as: 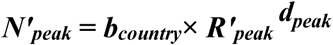.

## Results

### Dynamic changes in the reproduction rate

**Figure 2** shows the decreasing trends of observed ***R***_***d***_ for each country. As mentioned in the Methods section, ***R***_***d***_ is leveraged to characterize the overall reproduction rate in the past ***d*** days. Since ***R***_***d***_ is very dynamic in the first several days, we use the data of ***R***_***d***_ starting from day 12 to model the decreasing trend (as shown in the bottom of **Figure 2**). The coefficient β, intercept, adjusted r-squared, and p-values of β are shown in **Table 1**. Except for India, the linear regression models are learned at an average of adjusted r-squared as 0.96 and the 95% confidence interval as [0.94 0.98].

**Table 1.** Summary statistics of linear regression models to characterize the dynamic changes of the reproduction rate of COVID-19 case growth for each of the sixteen countries.

| Country | Coefficient $\beta$ | Intercept | Adjusted R-Squared | P-Value |
| --- | --- | --- | --- | --- |
| China | 0.0074946 | 1.4137 | 0.962 | 7.402e-21 |
| Korea | 0.0086331 | 1.4316 | 0.91 | 7.2914e-16 |
| Japan | 0.0010494 | 1.1159 | 0.889 | 3.9468e-15 |
| India | 0.00084686 | 1.1911 | 0.489 | 5.998e-05 |
| Iran | 0.0053851 | 1.3844 | 0.892 | 3.5508e-21 |
| Italy | 0.0043982 | 1.3642 | 0.987 | 1.4276e-38 |
| Germany | 0.0048399 | 1.383 | 0.993 | 4.4781e-41 |
| France | 0.0042749 | 1.3531 | 0.978 | 8.9805e-32 |
| Spain | 0.006088 | 1.4428 | 0.985 | 1.4977e-33 |
| Netherlands | 0.0041021 | 1.3065 | 0.992 | 4.0851e-34 |
| UK | 0.0032061 | 1.3121 | 0.987 | 3.862e-32 |
| USA | 0.0043401 | 1.4143 | 0.921 | 1.7286e-20 |
| Australia | 0.005486 | 1.3108 | 0.986 | 1.0694e-25 |
| Brazil | 0.0038629 | 1.2984 | 0.97 | 4.47e-20 |
| Argentina | 0.0045434 | 1.2382 | 0.966 | 4.1354e-14 |
| Egypt | 0.0012172 | 1.1417 | 0.923 | 1.5339e-14 |

**Figure 2.**
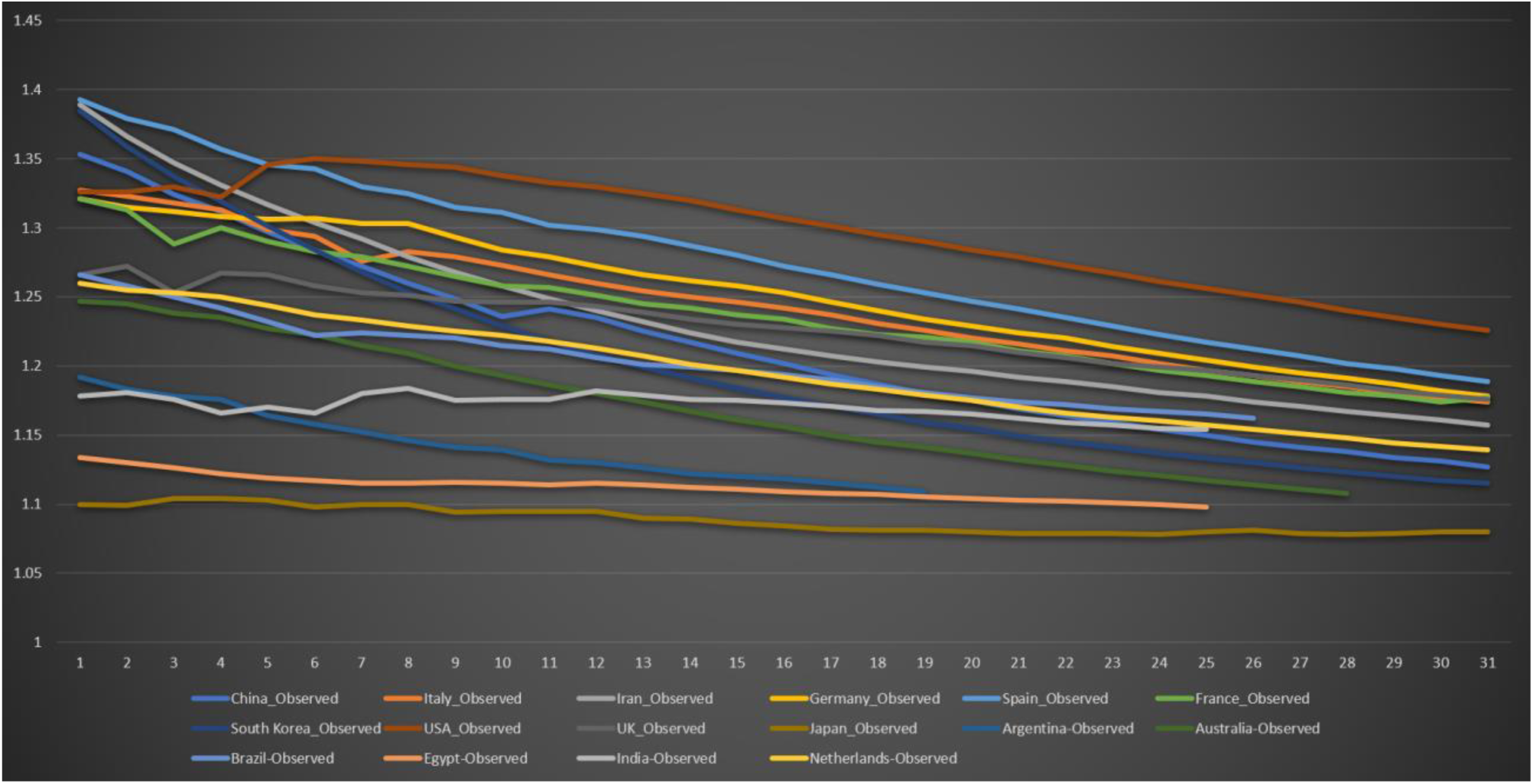
Decreasing trends of ***R***_***d***_ (the overall reproduction rate in the past ***d*** days since the 100^th^ case was confirmed) for each of the investigated countries. The data of ***R***_***d***_ starts from day 12 (index as 1 in the figure) since the 100^th^ case was confirmed.

### The cumulative number of cases conditioned by the day *d* and the overall reproduction rate in the past *d* days *R*_*d*_

**Figure 3** shows the comparative results of the observed cumulative number of COVID-19 cases (top) and the corresponding estimated number of cases conditioned by day ***d*** and the reproduction rate ***R***_***d***_ (bottom) among eight countries. The eight countries have the observed cumulative number of cases larger than 500 per million population by the time of the study (April 20, 2020). At the top of the figure, the number of observed cases (Y-axis) is normalized as the number of cases per million population. The X-axis is the number of days from the date when the 100^th^ case was confirmed. From the figure, it can be seen that Spain, Italy, and the US have both the highest observed and estimated cumulative number of cases per million population. Iran has the lowest observed and predicted number of cases. France, Netherlands, UK, and Germany are in the middle in terms of the observed and estimated cumulative number of cases.

**Figure 3.**
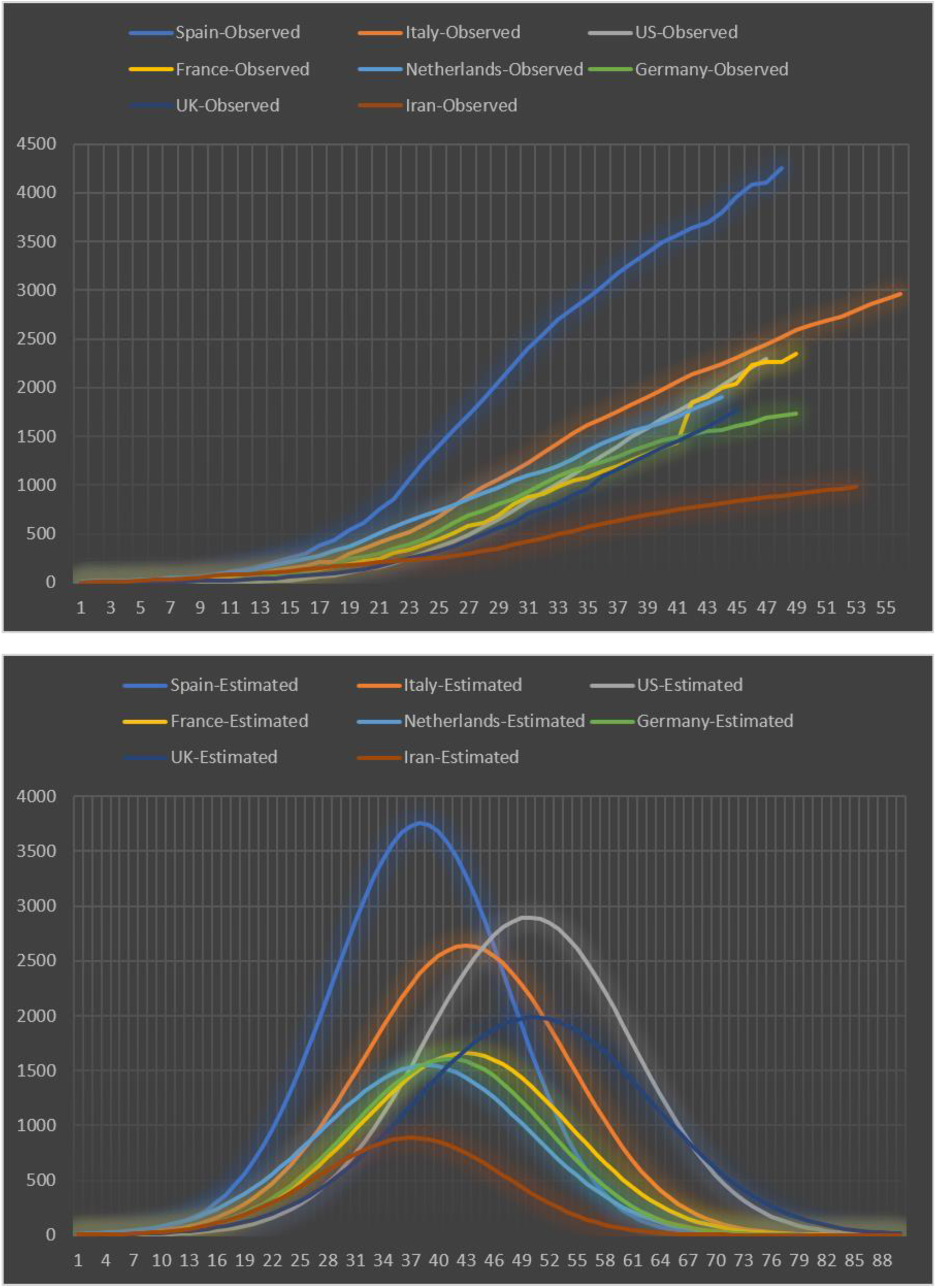
Growing trends of COVID-19 of the eight countries with the observed cumulative number of cases larger than 500 per million population (top) and corresponding estimated number of cases conditioned by day ***d*** and the reproduction rate ***R***_***d***_. The X-axis is the number of days ***d*** since the 100th case was confirmed, and the Y-axis is the cumulative number of the observed (top) and estimated (bottom) cases per million population.

Countries in **Figure 4** have smaller observed and predicted numbers of cases than the countries in **Figure 3**. Australia, South Korea, and Brazil have the largest number of the observed and estimated cumulative number of cases among the countries in Figure 4. Since the adjusted R-squared for the India model is less than 0.5 (**Table 1**), its estimated cumulative number of cases should not be considered in the analysis. The remaining four countries, including China, Japan, Egypt, and Argentina, have the smallest observed and estimated cumulative number of cases.

**Figure 4.**
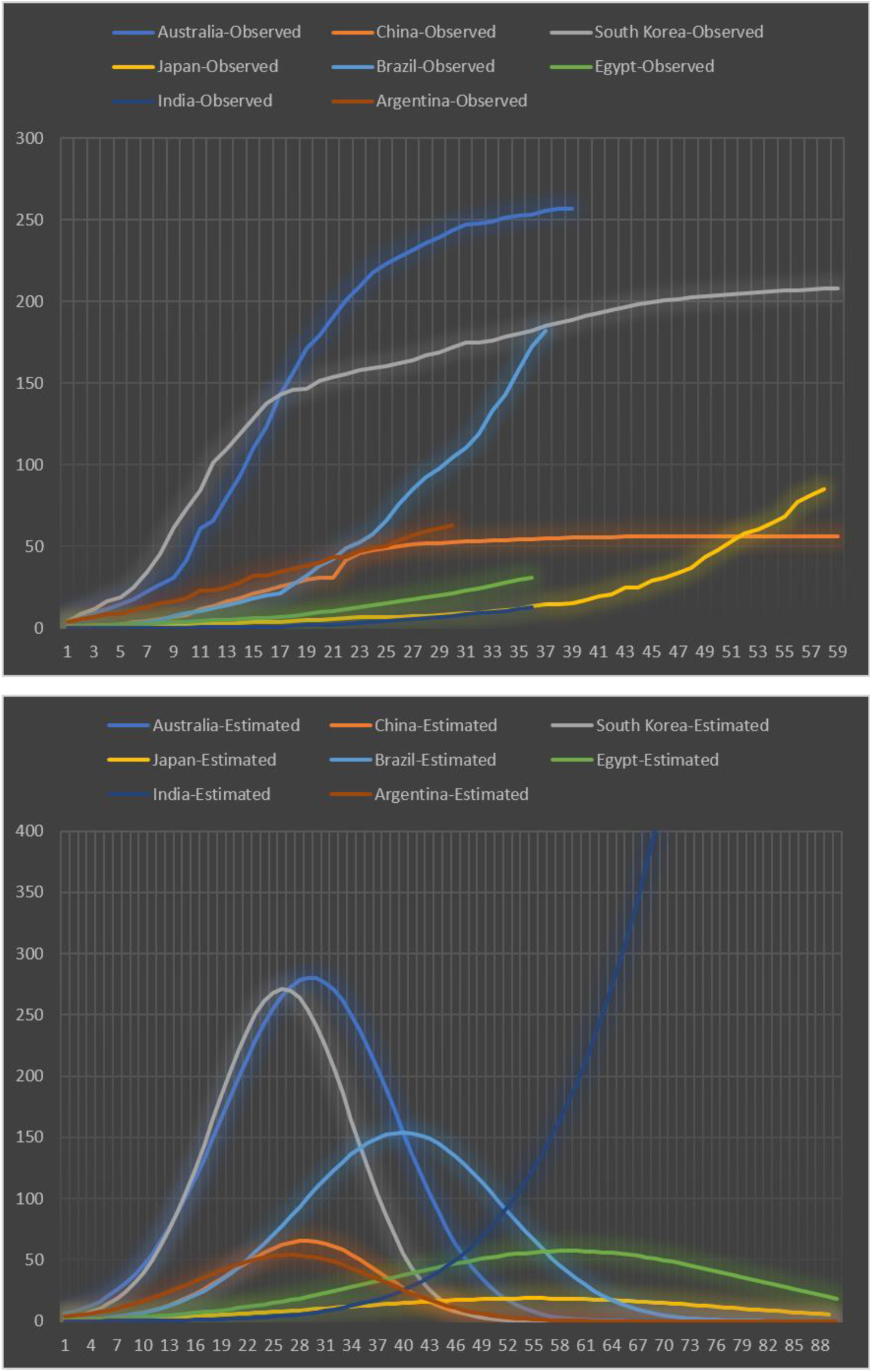
Growing trends of COVID-19 of the eight countries with the observed cumulative number of cases smaller than 500 per million population (top) and corresponding estimated number of cases conditioned by day ***d*** and the reproduction rate ***R***_***d***_. The X-axis is the number of days ***d*** since the 100th case was confirmed, and the Y-axis is the cumulative number of the observed (top) and estimated (bottom) cases per million population.

### Four groups of countries according to the effectiveness of their NPIs

**Figure 5** shows the four categories of countries according to the effectiveness of the NPIs. From the figure, it can be seen that the first class (the lower cumulative number and the smaller number of days) includes China, Argentina, Australia, South Korea, Brazil, and Iran. These countries have the most effective NPIs, and thus the overall reproduction rate is the lowest, and the number of days reaching the peak is the smallest. The second class includes Egypt and Japan. In the two countries, the cumulative number of cases is low at the peak, but it requires a long time to arrive at the peak (as shown in the bottom of **Figure 4**). The third group includes countries from Europe. Germany, Italy, Netherlands, Spain, and France have higher cumulative numbers of cases at the peak, but they require a relatively short time to reach the peak. The fourth class includes the US and UK, which have the highest cumulative number of fo cases at peak and require the longest time to reach the peak. It demonstrates the two countries have the most inefficient NPIs.

**Figure 5.**
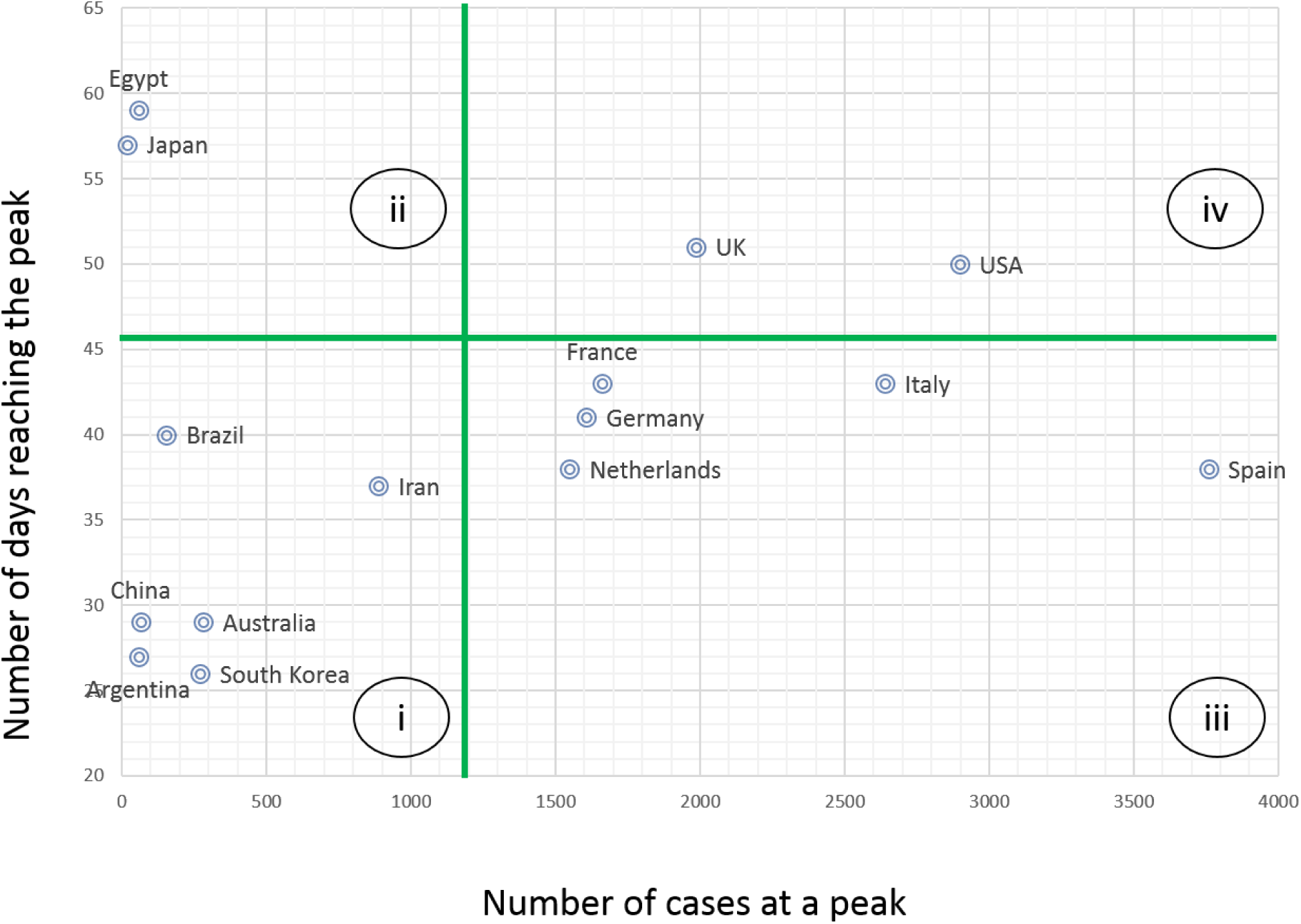
Groups of countries according to the number of days reaching the peak and the estimated cumulative number of cases at the peak.

## Discussion

It is very challenging to quantify the effectiveness of NPIs (e.g., social distancing, household quarantine) on COVID-19 spread trends in a direct way. In this paper, we use an indirect way to model the COVID-19 growth trend to reveal the differences in the effectiveness of NPIs among 16 representative countries. We leverage the dynamic change patterns of the reproduction rate to demonstrate the effectiveness of the NPIs. We categorize the investigated 16 countries into four groups according to the effectiveness of their adopted NPIs.

China and South Korea have the most effective NPIs. They are in the first group, as shown in **Figure 5**. Australia and Argentina are also included in the first group. Since the two countries are still at the beginning of the COVID-19 pandemic, potential subsequent outbreaks may change the reproduction rate and thus bring them into other groups. The first group uses the strategy to reduce the time to reach the peak and, at the same time, lower the height of the peak.

COVID-19 epidemic is a long way in Japan and Egypt. As shown in **Figures 4** and **5**, it still requires a longer time to arrive at their peaks. Fortunately, the estimated cumulative numbers of cases at the peaks are relatively small. The effectiveness of the NPIs in the two countries ranks second. Their strategy is to increase the time to reach the peak to avoid a higher one.

Countries in Europe are in the third class, where the effectiveness of the NPIs is in the media. The number of infected people per million population is very big for European countries. Except for the UK, all of the other investigated European countries almost arrive at their peaks. The US and UK are very special, and they are both in the fourth class, which has the most inefficient NPIs. They have the highest cumulative number of cases and, at the same time, require the longest time to arrive at their peaks.

The COVID-19 growth trend in India is hard to be modeled according to the existing data. There are several potential reasons. One of them could be the reported numbers are inaccurate, which could not represent the actual COVID-19 pandemic in India.

There are several limitations to this study that we wish to highlight to guide future research in this area. First, the dynamic changes in the reproduction rate may be impacted by many reasons other than the NPIs. If countries have other factors that influence COVID-19 growth, then our approaches to measuring the effectiveness of the NPIs will be limited.

Second, we use estimated peaks to categorize the effectiveness of the NPIs into four different levels. Although the estimated cumulative number of cases is consistent with the observed part (high adjusted R-Square), there is potentiality it will derive from the observed one in the future COVID-19 spread.

Third, the study aims to quantify the overall effects of NPIs on the case growing trend and did not consider the impact of each NPI, such as school and university close, on the growing trend. Due to limited data information and a lack of approaches, we did not separate each NPIs and consider all of them as a whole in this study.

Fourth, the patterns of COVID-19 growing are learned from reported case numbers, which may be underestimated given the shortages or unavailability of test kits in many countries. Findings learned from such data may be biased to the available number of test kits.

## Conclusions

We apply time-series COVID-19 case growth data to learn dynamic changes in the reproduction rate and leverage the reproduction rate as a tool to compare the effectiveness of the NPIs among countries in the world. We compared the effectiveness of NPIs for 16 countries. China, South Korea, Australia, and Argentina have the most effective NPIs, while the US and the UK have the most ineffective NPIs.

## Data Availability

We use publicly available data, which can be downloaded from https://data.humdata.org/dataset/novel-coronavirus-2019-ncov-cases

## Contributors

YC, YF, CY, XZ, and CG performed the data collection and analysis, methods design, experiments design, evaluation and interpretation of the experiments, and writing of the manuscript.

